# Bridging Acoustic and Semantic Spaces for Interpretable Voice Scoring via Zero-Shot Semantic Expansion

**DOI:** 10.64898/2026.05.29.26354442

**Authors:** Chi Hsiao, Yuan-Ren Cheng, Chung-Yao Yang, Fu-Shun Hsu

**Affiliations:** School of Medicine, Chung Shan Medical University, Taichung, Taiwan; Chung Shan Medical University Hospital, Taichung, Taiwan; Heroic Faith Medical Science Co., Ltd., New Taipei, Taiwan

**Keywords:** Voice pathology, Audio Spectrogram Transformer, Semantic alignment, Interpretable artificial intelligence, Zero-shot learning, Dysphonia, Objective voice assessment

## Abstract

Subjective auditory-perceptual evaluation and uninterpretable deep learning models limit the clinical assessment of voice disorders. This study proposes a two-phase zero-shot framework to evaluate voice pathology. First, an Audio Spectrogram Transformer is fine-tuned on the Perceptual Voice Quality Database to generate an acoustic latent space. Second, Orthogonal Procrustes analysis maps these acoustic embeddings directly onto the semantic space of a pre-trained Sentence Transformer. The geometric alignment produced continuous semantic axes that outperformed a supervised machine learning baseline in regressing clinician-rated GRBAS (Grade, Roughness, Breathiness, Asthenia, and Strain) severity scales. Furthermore, these axes correlate with traditional acoustic measures, including Harmonics-to-Noise Ratio and local jitter, while remaining robust when applied to aperiodic signals by not requiring fundamental frequency extraction. Most importantly, the model achieved zero-shot semantic expansion, successfully evaluating voices using an untrained, natural clinical vocabulary beyond the GRBAS scale. External validation on the Voice ICarus Database confirmed cross-corpus stability and demonstrated the capacity for zero-shot differential phenotyping of specific etiologies, such as hypokinetic dysphonia and reflux laryngitis. By bridging acoustic and semantic latent spaces, this framework offers an objective, continuous, and transparent metric for evaluating voice quality using voice descriptive vocabulary.

## 1. Introduction

Voice disorders are a common problem worldwide that presents with increasing prevalence since COVID [1]. Surveys of adults in the United States report lifetime prevalence rates of 20.6% to 29.9% [2, 3], where higher occurrence is found in the elderly and females [4]. Despite the burden, treatment-seeking rates remain low, with only 5.1% of affected individuals pursuing care [1]. Barriers include lack of awareness about available options, expectation of spontaneous resolution, and limited understanding of treatment purpose. Low perceived control over symptoms correlates with these barriers and a higher voice handicap [5, 6], suggesting that tools providing clear, easy-to-understand interpretation of pathologic audio may help patients grasp treatment benefits and motivate engagement. Clinically, assessment of dysphonia depends primarily on auditory-perceptual evaluation with scores such as GRBAS (Grade, Roughness, Breathiness, Asthenia, and Strain) and CAPE-V (Consensus auditory-perceptual evaluation of voice) [7], where each components is rated by experts on an ordinal scale. Acoustic analysis using Multi-Dimensional Voice Program (MDVP) [8] or Praat [9] may also be utilized to measure voice quality mathematically. For documentation of pathological voice, auditory-perceptual evaluation by clinicians is superior to acoustic measures with high validity and reliability, due to its ability to capture comprehensive properties of voices that acoustic measures fail to acknowledge [10, 11]. However, as results are influenced by listener experience and background, the intra-rater and inter-rater perceptual scoring of voices varies widely, sometimes receiving the full range of scoring for a single voice [12]. The need for a standard method to rate voices perceptually while avoiding the variability of human rating is valuable for further advances.

Deep learning models have been shown to distinguish pathological from healthy voices [13], approximate GRBAS rating [14], with high accuracy on curated datasets, though recent efforts also emphasize the need for model explainability in these domains [15]. While convolutional and recurrent architecture trained on dysphonic voice has achieved binary classification accuracies above 80%, transformer-based models offer significant advantages [16]. Models like the Conformer have demonstrated that self-attention mechanisms excel at capturing content-based global interactions and long-range dependencies [17]. Purely transformer-based models, such as the Audio Spectrogram Transformer (AST) [18], have increasingly been adapted to outperform traditional architectures in complex audio classification tasks. However, training a transformer requires a large amount of data, which is clinically difficult to acquire. When labeling is scarce, pre-trained transformer models via transfer learning provide foundational domain backbone structure while allowing flexibility for fine-tuning [19], and clinically, they have outperformed CNN baseline tasks such as classification of Parkinson’s disease diagnosis using voice data [20]. Yet, as seen in multi-class models [21], limiting the outputs of transformers only to diagnosis classification or ordinal categories compresses the continuous spectrum of dysphonia severity, which may not reflect the nuanced gradations perceived by clinicians. In addition, in small clinical datasets, deep learning models have been shown to passively learn shortcuts instead of genuine disease features [22], which, compounded by a lack of interpretability, leads to decreased trust and transparency [16, 23].

Traditional acoustic measures including jitter, shimmer, and harmonics-to-noise ratio (HNR) are clinically interpretable and widely used, but their validity depends on reliable pitch tracking.

Moreover, access to acoustic analysis requires software such as MDVP or Praat, which creates a barrier between the physician and the patient. According to signal typing frameworks, perturbation measures are appropriate only for Type 1 (nearly periodic) signals; for Type 2 (subharmonic) and Type 3 (chaotic) voices, pitch-detection algorithms fail, and the resulting jitter and shimmer values become unreliable or meaningless [24]. Cepstral peak prominence (CPP) does not require period detection and correlates well with overall dysphonia severity [25]. However, CPP is a single scalar that collapses roughness, breathiness, and strain into one value and does not equally capture all perceptual dimensions of the GRBAS scale [26]. Recent work on hybrid models that combine deep learning representations with low-level acoustic features has shown improved prediction of perceptual voice quality, suggesting that neither traditional acoustics nor deep learning alone is sufficient [27]. What remains missing is an objective, continuous metric that is robust to aperiodic signals, as deep learning embeddings are, while also being interpretable in terms of the semantic categories used to describe voice quality.

This study presents a two-phase framework for interpretable voice pathology evaluation. First, we fine-tune an Audio Spectrogram Transformer (AST) using clinical labels to generate an acoustic latent space. Second, rather than using standard regression outputs, we apply Orthogonal Procrustes Analysis to map these acoustic embeddings directly onto the semantic space of a pre-trained Sentence Transformer (Figure 1). We hypothesize this geometric alignment enables a zero-shot scoring system. Past literature demonstrates that pre-trained language representations can effectively capture the semantic similarity of clinical concepts without relying on manually curated domain-specific resources [28]. By linking acoustic data with clinical text, the model evaluates voice characteristics using descriptive vocabulary without requiring explicit supervised training on those specific terms.

**Figure 1.**
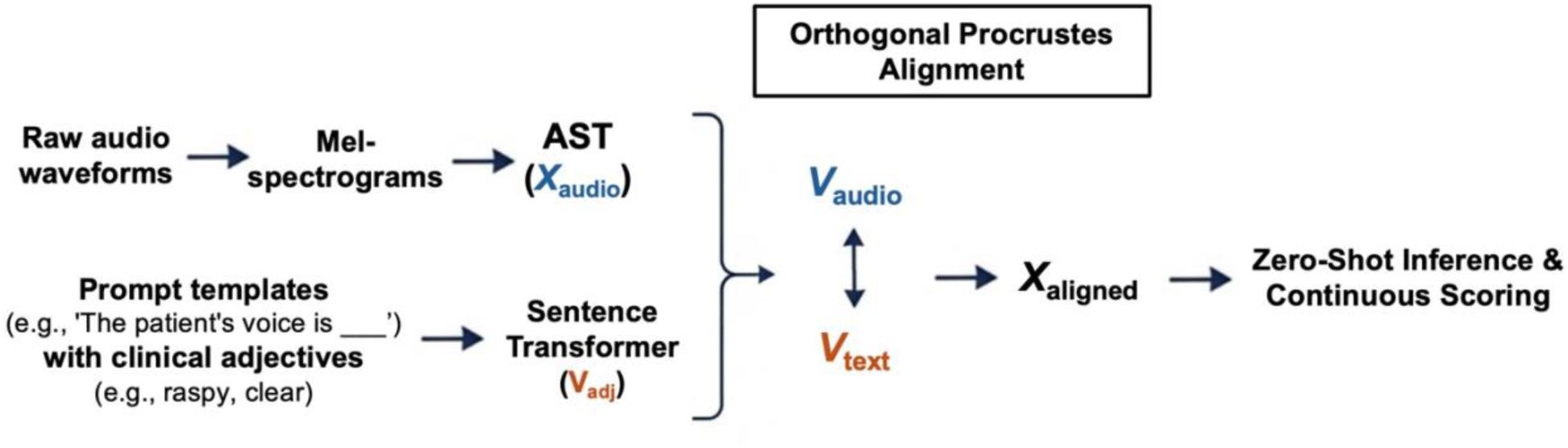
Overview of the zero-shot framework bridging acoustic and semantic latent spaces for interpretable voice pathology scoring. **Parallel Modality Extraction:** Raw audio waveforms are converted to mel-spectrograms and processed through a fine-tuned Audio Spectrogram Transformer (AST) to extract latent acoustic embeddings (𝑋_audio_). In parallel, clinical descriptive text is processed via a Sentence Transformer to generate semantic embeddings (𝑉_adj_). **Orthogonal Procrustes Alignment**: The entangled acoustic space is mathematically aligned with the structured semantic space. Acoustic anchors (𝑉_audio_) and text anchors are aligned using an optimal rotation matrix (𝑅), preserving the geometric relationships of the audio samples. Zero-Shot Inference and **Continuous Scoring**: Unseen patient audio (𝑋_test_) is projected into the aligned space (𝑋_aligned_). Clinical evaluation is performed by computing the cosine similarity between the aligned audio embedding and target clinical adjectives (𝐸_adj_), producing a continuous, multi-dimensional semantic voice profile.

## 2. Materials and methods

### 2.1. Datasets and Clinical Tables

The dataset used for model training and internal validation was the Perceptual Voice Quality Database (PVQD) [29]. The PVQD contains continuous speech samples of the "Rainbow Passage" collected from 296 participants. The demographic distribution includes 116 males and 180 females, with an average age of 50 years (range: 14 to 90). Clinical ground-truth labels for the PVQD samples were established using the GRBAS (Grade, Roughness, Breathiness, Asthenia, Strain) scale. Each parameter is scored on a discrete scale from 0 to 3, where 0 indicates an absent or normal condition and 3 indicates a severe condition. These labels were determined through an average of 3 expert clinicians ratings.

To evaluate the discriminative capability of the model, a subgroup analysis was conducted within the PVQD dataset. The clinical GRBAS Grade labels were used to establish distinct comparison groups: a healthy control group, defined by a Grade score threshold of <0.1, <0.25, or <0.5 (evaluated independently for threshold sensitivity), and a pathological group, defined by a Grade score equal to or greater than 1.0 or 2.0. For cross-corpus generalizability and zero-shot differential phenotyping, the Voice ICarus Database (VOICED) served as an external test set [30]. The VOICED dataset contains sustained vowel (/a/) phonations from 208 subjects, categorized into 57 healthy controls and 151 pathological cases. The pathological cohort is further classified by specific etiologies, including hyperkinetic dysphonia (n = 72), hypokinetic dysphonia (n = 41), and reflux laryngitis (n = 38). Unlike the PVQD, the VOICED dataset provides binary pathological classifications and specific etiological diagnoses rather than continuous perceptual GRBAS ratings.

### 2.2. Acoustic Feature Extraction and Parameter-Efficient Fine-Tuning

#### 2.2.1. Audio Preprocessing

All continuous speech samples were resampled to a target frequency of 16 kHz. Root Mean Square (RMS) normalization was applied to the raw audio waveforms. Mel-spectrograms were generated using a Short-Time Fourier Transform (STFT) with a window size of 1024, a hop length of 256, and 128 mel bins. The spectrograms were converted to a logarithmic decibel scale and standardized using global mean and standard deviation values computed from the training dataset. The temporal dimension of the inputs was fixed to 1024 frames. To meet the input requirements of the transformer model while accommodating varying audio lengths, zero-padding was applied to shorter samples. For longer samples, the cropping strategy depended on the modeling phase: random cropping was utilized during the training of the 5-fold cross-validation models to provide data augmentation, whereas center cropping was consistently applied when training the final model on the full dataset and during evaluation.

#### 2.2.2. AST Architecture and Parameter-Efficient Fine-Tuning

A pre-trained Audio Spectrogram Transformer (AST) (MIT/ast-finetuned-audioset-10-10-0.4593) was used to extract acoustic features. To optimize the latent space for voice pathology while preventing catastrophic forgetting of the pre-trained weights, the parameters of the base AST encoder were frozen except for the final encoder layer and the layer normalization parameters during fine-tuning. The model was trained to predict continuous GRBAS scores via ordinal regression using Mean Squared Error (MSE) loss. Optimization was performed using AdamW with a learning rate of 1 × 10^−4^, weight decay of 0.1, and a dropout probability of 0.3. For evaluation purposes, the continuous predictions were rounded to the nearest integer to map them to the discrete GRBAS categories. This configuration was first evaluated using stratified 5-fold cross-validation that reached a peak overall average weighted F1-score of 0.64 for predicting the discrete GRBAS categories, after which a final model was trained on the entire dataset using the same hyperparameters.

#### 2.2.3. Embedding Extraction

After fine-tuning the model on the clinical GRBAS labels, the regression output heads were discarded. The optimized 768-dimensional CLS token embeddings were extracted from the final encoder layer. These token embeddings served as the acoustic feature representations (𝑋_𝑎𝑢𝑑𝑖𝑜_) for the multimodal alignment phase.

#### 2.2.4. Traditional Acoustic Baseline

Standard acoustic metrics were extracted to physically validate the AI-derived semantic axes. Local Jitter, Local Shimmer, Harmonics-to-Noise Ratio (HNR), and Smoothed Cepstral Peak Prominence (CPPs) were computed using Praat via the Parselmouth Python librarys.

### 2.3. Semantic Latent Space Construction

Text embeddings were generated using the pre-trained all-mpnet-base-v2 model from the SentenceTransformers library. For the semantic anchors, clinical adjectives were embedded within a set of standardized prompt templates to provide context. Six templates were used, including phrases such as "The patient’s voice is " and "The audio demonstrates quality". Distinct sets of descriptive adjectives were defined to represent the "Severe" and "Normal" states for each GRBAS parameter. For the "Severe" state, specific clinical descriptors were used, such as "raspy" and "vocal fry" for Roughness, or "severely breathy" and "turbulent airflow" for Breathiness. For the "Normal" state, general descriptors including "normal," "healthy," "clear," and "stable" were applied.

### 2.4. Interpretable Metric Learning via Procrustes Alignment

Standard fine-tuning produces scalar predictions but lacks interpretability. To establish an explicit semantic basis, the optimized acoustic embedding space was mathematically aligned with the predefined clinical text embeddings using Orthogonal Procrustes analysis.

For each clinical parameter, the mean acoustic embeddings for severe cases (𝑉_severe_, defined as a score≥ 2) and normal cases (𝑉_normal_audio_ , defined as a score of 0) were calculated. The acoustic direction vector (𝑉_audio_) for each parameter was defined as the difference between these states (Eq. 1).

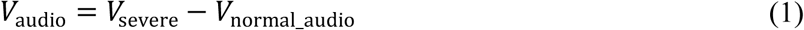

These parameter vectors constitute the audio anchor matrix. A corresponding text anchor matrix was generated using the language model embeddings for each target descriptive adjective (Eq. 2).

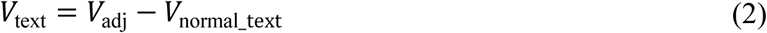

Then, Orthogonal Procrustes analysis was used to align the acoustic space with the semantic text space, calculating an optimal rotation matrix (𝑅) to map the audio anchors to the text anchors while preserving the geometric relationships between the audio samples. Unseen patient audio embeddings

(𝑋_test_) were mapped into the aligned semantic space (𝑋_aligned_ ) using this translation and rotation sequence (Eq. 3).

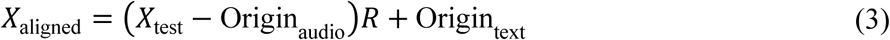

where Origin_audio_ is the mean vector of the normal voices, and Origin_text_ is the normal-state origin of the text space.

Clinical evaluation was conducted by measuring the angle between the aligned audio embedding (𝑋_aligned_) and a target text adjective embedding (𝑉_adj_) using cosine similarity (Eq. 4).

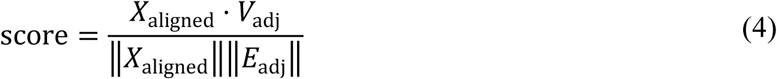

This calculation outputs a continuous value ranging from -1 to 1, representing the proximity of the acoustic sample to the specified clinical descriptor. To standardize these values for clinical interpretation, the cosine similarity scores were normalized into Z-scores based on the statistical distribution of the normal (Grade 0) reference group.

### 2.5. Statistical Analysis and Experimental Design

To evaluate the system and prevent overfitting, a 5-fold cross-validation scheme was implemented on the PVQD dataset. The performance was assessed by categorizing voices as Normal or Pathological. The proposed Procrustes alignment method was benchmarked against a supervised baseline. This baseline consisted of a Logistic Regression classifier, optimized with Principal Component Analysis (PCA) for dimensionality reduction, trained directly on the AST CLS tokens. Classification performance was quantified and compared using the F1-score and the Area Under the Receiver Operating Characteristic Curve (AUC-ROC). Convergent validity was assessed using Spearman’s rank correlation coefficient (𝑟_s_) to compare Procrustes cosine similarity scores with standard Praat acoustic features.

## 3. Results

### 3.1. Geometric Disentanglement of Acoustic and Semantic Spaces

The Orthogonal Procrustes alignment successfully mapped the latent acoustic representations of the fine-tuned AST onto the target semantic space. Prior to alignment, the acoustic vectors corresponding to the GRBAS parameters were entangled within the continuous latent space.

Following the application of the rotation matrix (𝑅), the latent space geometrically separated into distinct, interpretable semantic axes (Figure 2). The proposed Procrustes alignment method was benchmarked against a supervised baseline trained on the unaligned AST CLS tokens (Table 1). The Procrustes alignment method demonstrated superior performance across all five GRBAS parameters in terms of both F1-score and AUC-ROC. This confirms that mathematically aligning the acoustic and semantic spaces not only provides interpretability but also enhances the capacity to reliably stratify normal versus pathological voices.

**Figure 2.**
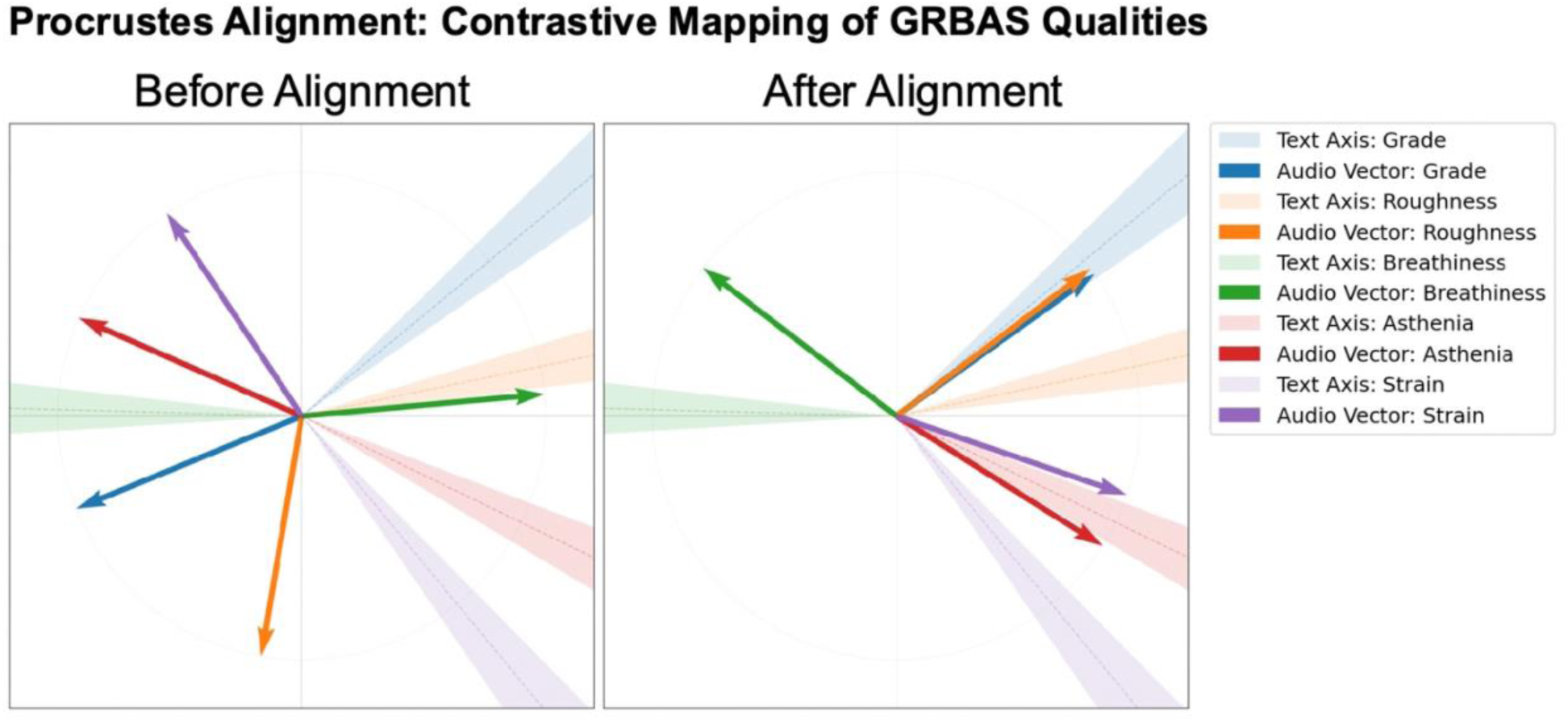
Contrastive Principal Component Analysis (PCA) of the acoustic latent space. The plot presents the distribution of acoustic (arrows) and text (rays) embedding anchors before and after Orthogonal Procrustes alignment. Unaligned embeddings demonstrate entanglement across pathological features, while post-alignment embeddings separate into distinct geometric trajectories closer to the GRBAS clinical dimensions.

**Table 1.** Method comparison between the Supervised Baseline and Procrustes Alignment on the Perceptual Voice Quality Database (PVQD). Performance is evaluated for binary classification (Normal < 0.1 vs. Pathological ≥ 2.0) across all GRBAS parameters using F1-score and Area Under the Curve (AUC). Positive improvement values indicate the superior discriminative capacity of the Procrustes-aligned semantic space compared to a supervised logistic regression model trained on unaligned acoustic embeddings.

|  | CLS Base |  | Our method |  | Improvement |  |
| --- | --- | --- | --- | --- | --- | --- |
| Parameter | F1 | AUC | F1 | AUC | F1 | AUC |
| Grade | 0.36 | 0.32 | 0.72 | 0.68 | +0.36 | +0.35 |
| Roughness | 0.21 | 0.10 | 0.62 | 0.84 | +0.41 | +0.74 |
| Breathiness | 0.41 | 0.62 | 0.60 | 0.85 | +0.18 | +0.23 |
| Asthenia | 0.19 | 0.28 | 0.50 | 0.81 | +0.31 | +0.53 |
| Strain | 0.54 | 0.39 | 0.58 | 0.57 | +0.04 | +0.18 |

To ensure the Procrustes alignment was not driven by the syntactic structure of the prompt templates, a sensitivity analysis was conducted using external semantic anchors (Supplementary Tables 1 and 2). The absolute axis trajectories maintained high directional consistency with an average cosine similarity > 0.86. More importantly, the inter-parameter geometric structure was preserved across prompt variations (Spearman’s 𝜌 ranging from 0.830 to 0.976), indicating that the derived semantic space captures the core clinical adjectives rather than superficial syntactic patterns.

**Table 2.** Spearman correlation coefficients between the semantic axes and traditional acoustic parameters. Acoustic metrics were extracted via Praat. The data validate the physical grounding of the semantic space in established acoustic phenomena, including perturbation and aerodynamic noise.

| Procrustes Semantic Axis | Acoustic Correlate (Praat Measure) | Spearman's Correlation (rs) | p-value |
| --- | --- | --- | --- |
| Roughness | Local Jitter (%) | 0.45 | <0.001 |
| Breathiness | HNR (dB) | -0.58 | <0.001 |
| Asthenia | F0 Standard Deviation (Hz) | 0.24 | <0.001 |
| Strain | Local Shimmer (dB) | 0.39 | <0.001 |

### 3.2. Alignment with Human Clinical Perception

To evaluate whether the semantic scores correspond to human auditory perception, they were compared against the ordinal clinical severity gradings (Figure 3). As the clinician-rated severity increases, the median continuous Procrustes Z-score reliably and monotonically shifts upward across all five parameters, confirming that the mathematically derived semantic axes for Grade, Roughness, Breathiness, Asthenia, and Strain accurately map to the perceptual severity graded by human experts. The distributions indicate that the continuous scoring system preserves the gradations of clinical severity while providing a higher-resolution measurement than standard discrete integers.

**Figure 3.**
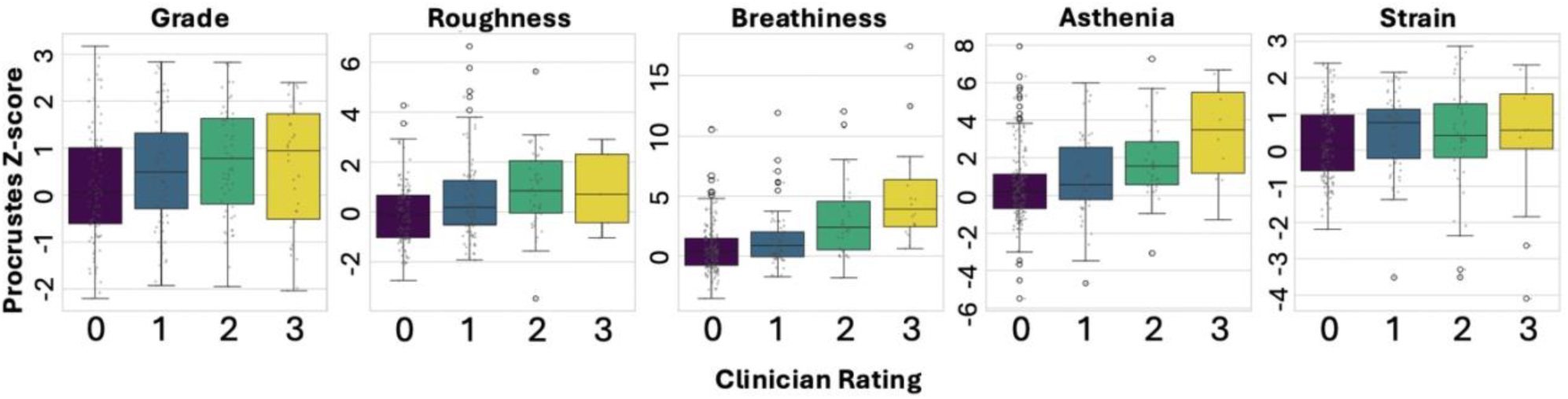
Distribution of continuous Procrustes semantic scores across ordinal clinical severity grades. Boxplots demonstrate the normalized continuous Z-scores for Grade, Roughness, Breathiness, Asthenia, and Strain plotted against the ground-truth clinician ratings (0 to 3). The positive monotonic shift indicates alignment between the mathematically derived semantic axes and human auditory perception. The boxplot displays the median, the 25th and 75th percentiles (box), and the 1.5×interquartile range (whiskers); individual points indicate outliers.

### 3.3. Zero-Shot Semantic Expansion and Construct Validity

After the Orthogonal Procrustes method aligns the entire latent acoustic space with the continuous semantic space of the language model, we assume the system has acquired the capacity to evaluate clinical vocabulary beyond the explicit training labels. The construct validity heatmap reveals strong positive correlations with established synonyms for specific pathologies (Figure 4); for example, the Roughness axis is positively associated with the descriptor "raspy." Conversely, strong negative correlations are observed with antonyms, such as the inverse relationship between the Grade axis and the descriptor "clear." This structural alignment indicates that the model generalizes to related clinical terminology without requiring explicit training on those specific words.

**Figure 4.**
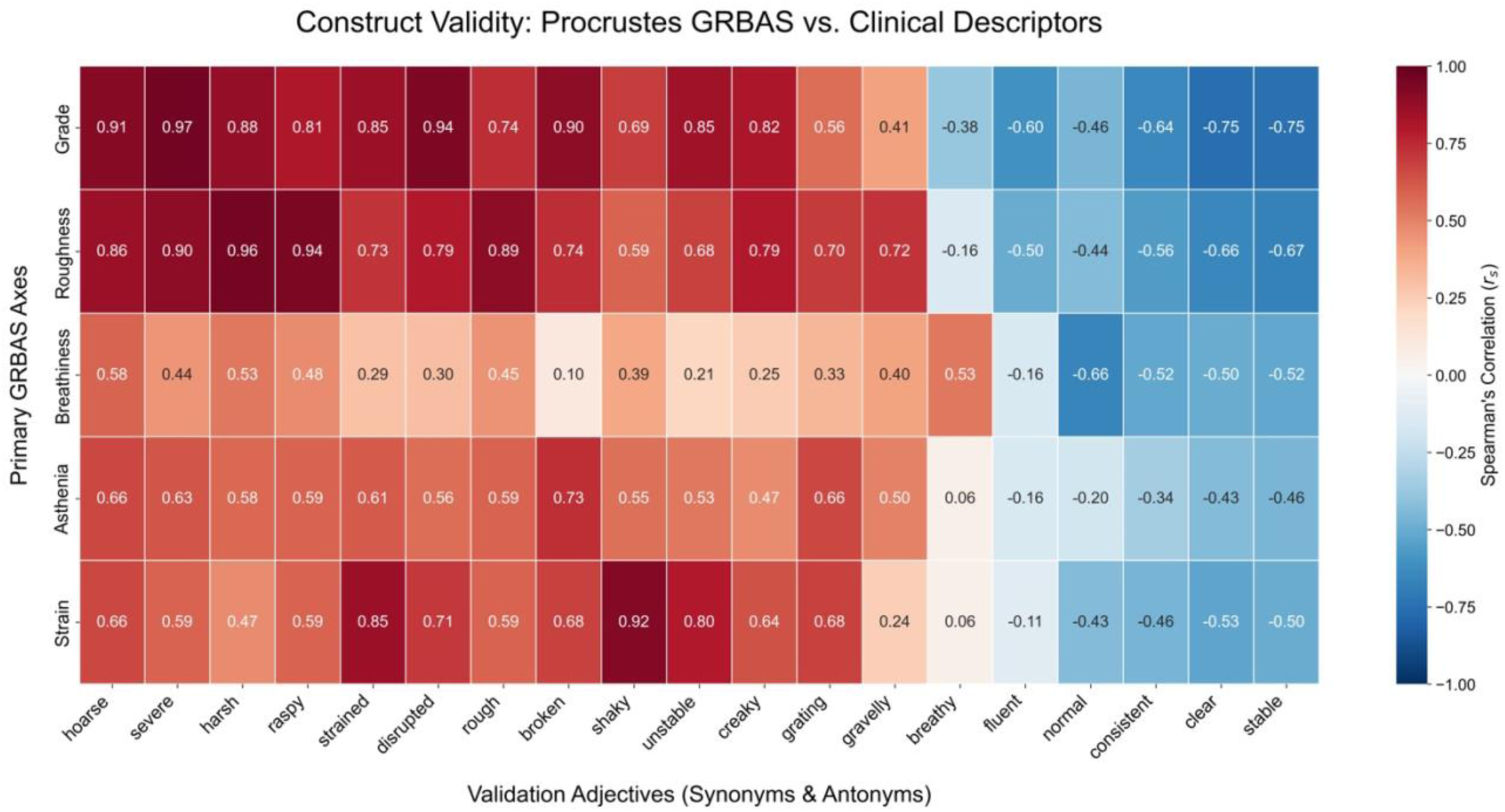
Construct validity heatmap of zero-shot semantic expansion. The heatmap illustrates the correlation between the aligned acoustic semantic axes and an expanded vocabulary of clinical descriptors. Positive correlations are observed with established synonyms (e.g., Roughness and "raspy"), and negative correlations are observed with antonyms (e.g., Grade and "clear").

To evaluate the clinical utility of this multidimensional expansion, the model generated semantic voice profiles for distinct patient archetypes (Figure 5). Traditional acoustic metrics lack clear differentiation between high roughness and high strain, as single-scalar measures like Cepstral Peak Prominence (CPP) combine these perceptual dimensions into one value. Our method of aligned semantic adjectives successfully separates these two voice qualities. By projecting the acoustic embeddings onto specific descriptive terms, the model provides a distinct, multidimensional profile that differentiates high roughness from high strain, addressing a known limitation of standard acoustic analysis.

**Figure 5.**
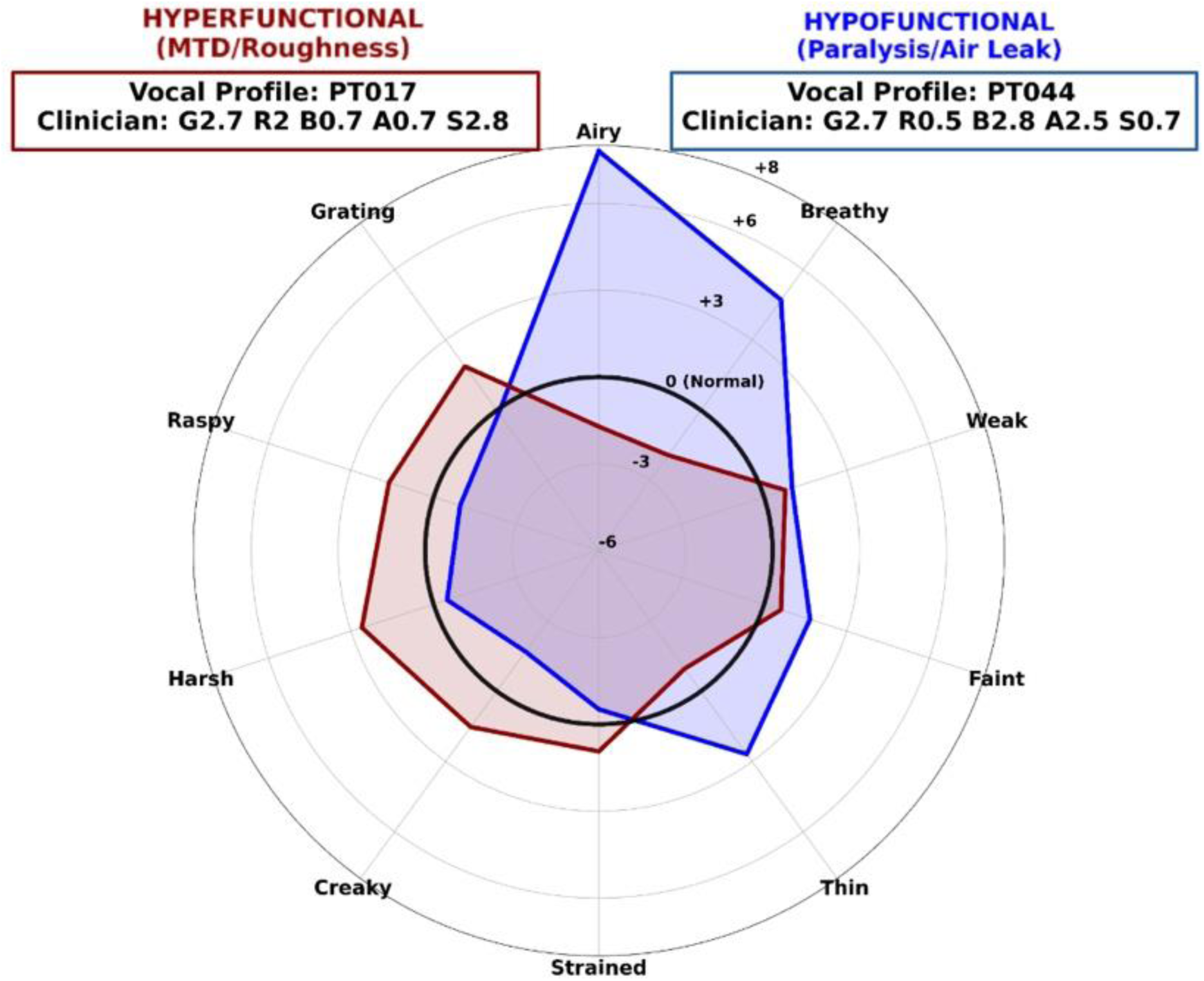
Multidimensional semantic voice profiles for distinct patient archetypes. Radar charts map individual acoustic samples onto specific descriptive adjectives. The derived high strain and high roughness profiles demonstrate the ability of the model to geometrically differentiate complex voice qualities, which is limited in traditional single-scalar acoustic metrics.

### 3.4. Physical Grounding in Traditional Acoustics

To validate that the mathematically derived semantic space learned physical acoustic properties, the continuous semantic axes were compared against traditional acoustic measures extracted via Praat (Table 2). The Breathiness axis demonstrated a significant negative correlation with the Harmonics-to-Noise Ratio (HNR) (𝑟_𝑠_ = -0.5829), while the Roughness and Strain axes had significant positive correlations with local jitter (𝑟_𝑠_ = 0.4453) and local shimmer (𝑟_𝑠_ = 0.3942), indicating that the model successfully captured aerodynamic noise and acoustic perturbation within the voice signal.

Consequently, the model evaluates these physical phenomena without requiring fundamental frequency (𝐹_0_) extraction. This allows the system to function on aperiodic or chaotic signals where traditional pitch-detection algorithms fail and acoustic perturbation measures become unreliable.

### 3.5. Baseline Severity Detection and Zero-Shot Etiological Profiling

The diagnostic capacity of the Procrustes Grade metric was evaluated on both the unseen PVQD test folds and the external VOICED dataset (Table 3). On the PVQD cohort, the model demonstrated sensitivity to the clinical definition of normophonia. When the healthy control group was defined broadly to include mild deviations (Grade < 0.50), the AUC for detecting moderate-to-severe pathology (Grade ≥ 2.0, n = 62) was 0.62. When the control threshold was restricted to voices with near-zero perceptual deviation (Grade < 0.10, n = 31), the AUC increased to 0.71, indicating the metric’s responsiveness to sub-clinical acoustic shifts. Upon external validation using the VOICED dataset, the Grade metric achieved an AUC of 0.65 for all diseased patients.

**Table 3.** Zero-Shot Baseline Severity (’Grade’) Performance Across Corpora. Area Under the Receiver Operating Characteristic Curve (AUC-ROC) for the continuous Procrustes ’Grade’ metric. For the internal PVQD test folds, performance is evaluated across varying definitions of the normophonic control group (from Grade < 0.50 to Grade < 0.10) against moderate-to-severe pathology (Grade > 2.0). For the external VOICED dataset, performance is evaluated for all pathological samples and isolated etiologies against healthy controls. N represents sample count.

| Dataset | Control | Pathology | N<br>(Control / Path) | Grade<br>AUC |
| --- | --- | --- | --- | --- |
| **PVQD**<br>**(Holdout)** | Clinical Grade < 0.50 | Grade > 2.0 | 77 / 62 | 0.62 |
|  | Clinical Grade < 0.25 | Grade > 2.0 | 52 / 62 | 0.65 |
|  | Clinical Grade < 0.10 | Grade > 2.0 | 31 / 62 | 0.71 |
| **VOICED**<br>**(External)** | Healthy (n=57) | All Diseased (n=151) | 57 / 151 | 0.65 |
|  | Healthy | Hypokinetic Dysphonia (n=72) | 57 / 41 | 0.68 |
|  | Healthy | Hyperkinetic Dysphonia (n=41) | 57 / 72 | 0.65 |
|  | Healthy | Reflux Laryngitis (n=38) | 57 / 38 | 0.62 |

Performance varied slightly across specific etiologies, yielding AUCs of 0.68 for Hypokinetic Dysphonia, 0.65 for Hyperkinetic Dysphonia, and 0.62 for Reflux Laryngitis. These results suggest that while a unidimensional severity score detects general acoustic degradation, it does not fully separate distinct pathophysiological states.

To evaluate the model’s capacity for differential phenotyping, zero-shot semantic profiling was conducted on the VOICED disease cohorts using the expanded adjective space (Table 4). The multi-dimensional semantic distances aligned with established clinical pathophysiology. For Hypokinetic Dysphonia, a condition characterized by vocal fold bowing and reduced respiratory drive, the model identified "weak" (AUC = 0.810), "faint" (AUC = 0.806), and "asthenic" (AUC = 0.803) as the strongest semantic predictors. Conversely, for Reflux Laryngitis, characterized by edematous vocal folds, the model yielded inverse predictive relationships for "breathy" (AUC = 0.342) and "airy" (AUC = 0.340), correctly predicting a lack of glottic insufficiency. Hyperkinetic Dysphonia was best characterized by "muffled" (AUC = 0.663) and "hoarse" (AUC = 0.650). These findings indicate that the Procrustes-aligned latent space preserves the geometric signatures required to differentiate specific vocal etiologies without explicit supervised training.

**Table 4.** Zero-Shot Semantic Phenotyping of Specific Etiologies of VOICED. Top discriminative semantic features for distinct pathological cohorts. AUC-ROC values represent the capacity of the zero-shot semantic distance to distinguish the specific etiology from healthy controls. Values below 0.50 (*) indicate an inverse relationship, wherein the pathological cohort scores lower on the respective semantic axis than the healthy controls. Effective discriminative power for inverse relationships is derived as (1 - AUC).

| Disease | Semantic Feature | AUC-ROC | Clinical Relationship | Pathophysiological Interpretation |
| --- | --- | --- | --- | --- |
| **Hypokinetic Dysphonia** | Weak | 0.81 | Positive | Reflects reduced respiratory drive / vocal fold bowing. |
|  | Faint | 0.806 | Positive | Diminished vocal intensity. |
|  | Asthenic | 0.803 | Positive | Classic GRBAS parameter for hypofunction. |
|  | Low-pitched | 0.803 | Positive | Loss of dynamic pitch range. |
| **Reflux Laryngitis** | Breathy | 0.342<br>(0.658)* | Inverse | Heavy, edematous folds prevent air leakage. |
|  | Airy | 0.340<br>(0.660)* | Inverse | Opposite of glottic insufficiency. |
|  | Strong | 0.335<br>(0.665)* | Inverse | Vocal fatigue limits projection. |
| **Hyperkinetic Dysphonia** | Muffled | 0.663 | Positive | Over-compression of the vocal tract. |
|  | Hoarse | 0.65 | Positive | Aperiodic vocal fold vibration due to strain. |

## 4. Discussion

Current deep learning models in voice pathology assessment often prioritize classification accuracy at the expense of clinical interpretability and transparency [31–36]. To address this, our proposed framework geometrically aligns the latent acoustic representations of an Audio Spectrogram Transformer (AST) with the semantic space of a large language model [37], backed by the alignment hypothesis [38]. This approach generates a continuous, multi-dimensional metric, allowing physicians to see exactly which semantic characteristics and clinical descriptors contributed to the diagnostic evaluation [27, 39, 40]. Most importantly, aligning these complete latent spaces enables zero-shot semantic expansion [41], as the comparable embeddings could be generalized to related clinical vocabulary, evaluating unseen adjectives without requiring explicit supervised training [28, 42, 43]. Consequently, the system allows the model to capture complex voice qualities such as severe roughness and strain (Figure 4 and 5).

Traditional acoustic metrics like jitter and shimmer are the standard for objective voice evaluation, but they struggle with severely dysphonic voices [44, 45]. Because these perturbation measures rely on fundamental frequency (𝐹_0_) extraction, they frequently fail or become unreliable when applied to aperiodic or chaotic signals (Type 2 and Type 3 voices) [46, 47]. Our AST-based framework bypasses this limitation entirely because it does not require 𝐹_0_ extraction to generate its latent representations [18]. Importantly, despite lacking pitch tracking, the derived semantic space remains physically grounded. As shown in Table 2, our mathematically derived Breathiness axis significantly correlates with the Harmonics-to-Noise Ratio (HNR). This demonstrates that the model successfully captures the physical principles of vocal aerodynamics and noise while remaining robust to severe signal degradation.

A critical finding of this study relates to the sensitivity of the model to the clinical definition of “normal” (Table 3). When the healthy control group was strictly limited to voices with near-zero perceptual deviation (Grade < 0.10), the zero-shot alignment achieved an AUC-ROC of 0.71 against the moderate-to-severe pathology group. However, when this baseline was broadened to include voices with mild dysphonia (Grade < 0.50), the classification performance progressively dropped to 0.62. This pattern demonstrates that the continuous Procrustes metric is highly sensitive to micro-acoustic deviations within the "normal" spectrum. A human evaluator might functionally categorize a Grade 0.4 voice as healthy for clinical triage, but the geometric alignment detects that its acoustic embedding has already begun drifting toward the pathological semantic space [48].

Ultimately, this indicates that the continuous metric evaluates acoustic degradation at a higher resolution than ordinal human labels, successfully detecting sub-clinical variations before they cross the subjective threshold of perceived pathology.

Certain limitations in the current study must be acknowledged. First, the predictive accuracy of the proposed framework is lower than that of a standard supervised deep learning model [21]. However, the proposed framework sacrifices some predictive power to establish a transparent, continuous evaluation scale [49, 50], while avoiding overfitting on a small clinical dataset [51]. The semantic alignment itself also provides distinct structural benefits. When benchmarked against a linear baseline trained directly on unaligned acoustic embeddings (Table 1), the zero-shot semantic mapping demonstrated consistent improvements in F1-scores and AUC-ROC. This indicates that while the system may not match the absolute accuracy of fully supervised, task-specific models, grounding the acoustic latent space in a structured linguistic model actively enhances feature separability. Second, the internal validation relied on the Perceptual Voice Quality Database (PVQD), which utilizes human-generated GRBAS ratings as the reference standard. Because auditory-perceptual evaluation is inherently subjective, the baseline labels contain inherent human variability and cross-rater noise [52]. This subjectivity places a mathematical upper limit on the achievable accuracy of any objective metric evaluated against these human scores.

To evaluate cross-corpus generalizability, the model was tested on the external VOICED dataset[30]. Despite a structural domain shift (training on continuous speech but evaluating on sustained vowels (/a/)), the baseline severity metric maintained stable performance for detecting diseased patients. More importantly, this external validation demonstrated the ultimate clinical utility of zero-shot semantic expansion (Table 4). Without any explicit training on specific etiologies, the framework successfully performed differential phenotyping. It accurately characterized hypokinetic dysphonia using descriptors like "weak" and "faint," and correctly predicted the absence of glottic insufficiency ("airy," "breathy") in reflux laryngitis. These findings confirm that the geometrically aligned semantic space preserves the critical structural signatures needed to differentiate complex pathophysiological states across diverse acoustic tasks and clinical populations.

The method presented in this article if highly valuable for tracking multi-dimensional acoustic changes during vocal rehabilitation or following surgery, as it has the capacity to move voice assessment beyond the rigid constraints of the GRBAS scale. In conclusion, geometrically aligning acoustic features with a semantic language model successfully resolves the interpretability limitations of standard deep learning. By achieving zero-shot semantic expansion, this framework empowers clinicians to evaluate and document voice pathology using an expansive, natural descriptive vocabulary. Ultimately, this provides a highly transparent and clinically intuitive tool that bridges advanced computational acoustics with real-world laryngology.

## 5. Data and Code Availability Statement

The datasets analyzed during the current study are available in public repositories. The Perceptual Voice Quality Database (PVQD) used for internal training and validation is accessible at https://data.mendeley.com/datasets/9dz247gnyb/4. The external validation dataset, Voice Icarus Database (VOICED), is available at https://physionet.org/content/voiced/1.0.0/.

The custom Python code and the Orthogonal Procrustes alignment matrices required to reproduce the core findings of this study are publicly available on GitHub at https://github.com/amyhsiao/SemanticVoice/. The repository includes a minimal reproducible example demonstrating the zero-shot semantic expansion pipeline on sample audio.

## 6. Authors’ contributions

**Chi Hsiao:** Conceptualization, Methodology, Formal analysis, Writing - original draft, Writing - review & editing. **Yuan-Ren Cheng:** Conceptualization. **Chung-Yao Yang:** Conceptualization. **Fu-Shun Hsu:** Conceptualization, Supervision. All authors read and approved the final manuscript.

## 8. Declaration of generative AI use

During the preparation of this work the author(s) used Gemini 3.0 to refine the English phrasing, improve readability, and organize the structural flow of the manuscript. After using the service, the author(s) reviewed and edited the content as needed and take(s) full responsibility for the content of the published article.

## Data Availability

All data produced in the present work are contained in the manuscript.

https://github.com/amyhsiao/SemanticVoice/

