## Supplementary material for "Bridging Acoustic and Semantic Spaces for Interpretable Voice Scoring via Zero-Shot Semantic Expansion": Supp

**Supplementary Table 1. Prompt sensitivity and semantic space stability.** Comparison of average cosine similarity, structural correlation (Spearman's ρ), and origin shift (L2) across alternative prompt sets (Casual, Formal Medical, Minimal) to confirm that the inter-parameter geometric structure is preserved independent of syntactic variation.

| Prompt Set | Avg Cosine Similarity | Structural Correlation (rho) | Origin Shift (L2) |
|---|---|---|---|
| **Casual (The speaker sounds...)** | 0.887 | 0.83 | 0.517 |
| **Formal Medical** | 0.864 | 0.976 | 0.527 |
| **Minimal (The voice is...)** | 0.918 | 0.927 | 0.455 |

**Supplementary Table 2: Catalog of semantic prompt templates.** The comprehensive list of standardized syntactic templates used to embed clinical adjectives and construct the target semantic space for zero-shot alignment.

| Prompt Category | Template | Purpose |
|---|---|---|
| **Original Baseline** | The patient's voice is {}. | Primary semantic space alignment |
| | The audio demonstrates {} quality. | Primary semantic space alignment |
| | A {} voice. | Primary semantic space alignment |
| | Auditory perception: {}. | Primary semantic space alignment |
| | The voice sounds {}. | Primary semantic space alignment |
| | Evidence of {} in the vocal signal. | Primary semantic space alignment |
| **Casual** | The speaker sounds {}. | Sensitivity analysis for syntactic variance |
| **Formal Medical** | Clinical assessment confirms a {} vocal presentation. | Sensitivity analysis for syntactic variance |
| | Pathological analysis identifies {} characteristics. | Sensitivity analysis for syntactic variance |
| | The larynx produces a {} sound. | Sensitivity analysis for syntactic variance |
| **Minimal** | The voice is {}. | Sensitivity analysis for syntactic variance |
